# The age and sex distribution of COVID-19 cases and fatalities in India

**DOI:** 10.1101/2020.07.14.20153957

**Authors:** Sourendu Gupta

## Abstract

Using anonymous publicly available data on COVID-19 infections and gross outcomes in India, the age and sex distribution of infections and fatalities is studied. The age structure in the count of infections is not proportional to that in the population, indicating the role of either co-morbidity or differential attack rate. There is a strong age structure in the sex ratio of cases, with the female to male ratio being about 50% on average. The ratio drops between puberty and menopause. No such structure is visible in the sex ratio of fatalities. The overall age distribution of fatalities is consistent with a model which uses the empirical age structure of infections and a previous determinations of age structured IFR. The average IFR for India is then expected to be 0.4% with a 95% CrI in [0.22%, 0.77%].

## I. INTRODUCTION

The chance of surviving an infectious disease is not an absolute number. It depends on medical advances, and the level of care available to patients. The former is significant as the COVID-19 pandemic progresses, with new drugs [1] and treatment regimes [2] being developed. The level of medical care available also depends on economic and regional factors [3], as well as policy matters like the amount of protection and testing given to health workers. Nevertheless, following an early attempt to predict the burden of COVID-19 in France [4], there have been attempts to extend this to other geographical regions of the world [5] including India [6, 7].

Infection by SARS-CoV-2 results in a diverse set of pathologies [8] with disruptive outcomes. The infection fatality ratio, IFR, is the probability that an infection results in a fatality. IFR can be empirically extracted from the ratio of deaths to all infections in any sufficiently large random sample of the population. In an ongoing epidemic this is hard to estimate, especially when testing is limited. The widely used alternative, case fatality ratio, which is the ratio of fatality to all test positives, depends on the number of tests administered and the policy for administering tests [9]. It is a biased estimator of the probability of survival following infection.

A priori estimation of IFR requires a large random sample of infected persons and intensive longitudinal records of testing and treatment, in order to determine the chances of recovery. All the projections mentioned before took as input an analysis in [10], which extracted the age structure of IFR from a variety of sources, with an integrated sample size of 44672. The age-stratified tables of IFR from [10] were converted to a population averaged value for India in [7] using the age distribution of the population according to the 2011 census of India [11]. There are three assumptions made in this estimate. The first is that the age distribution of infections is the same as that of the population. The second is that the pattern of co-morbidities, which are important for the severity of COVID-19, is similar to that intrinsically in the work of [10]. The final assumption is that genetic factors are negligible.

The first two assumptions are clearly simplifying hypothesis which are now open to test. That genetics is important in the progression of the pathology of COVID-19 infections, including fatalities, is becoming clear. Studies have reported that the severity of the disease [12] as well as the mortality burden [13] among women is smaller. There has been suggestions about the link between these observations and X-chromosome heterozygosity in women [14]. Some correlations between the disease and the ABO blood group, as well as other genetic loci, have been reported [15].

The publicly available crowd-sourced data [16] for India is used to examine various aspects of the epidemiology of COVID-19 infections in India using aggregate Indian data. Significant age-structured sex-differences in the number of infections is observed. The age-structures sex-differences in the number of fatalities is observed to differ quite significantly. This observation calls for further follow up using clinical data, and alternate data sets. An age and sex averaged IFR for India is predicted. The lack of further clinical details in the public domain is an impediment to asking other useful questions. Among these are clinically important issues such as the time intervals between different stages in the progress of the disease, which is needed to estimate the public-health infrastructure which needs to be made available. Also important is the issue of long-term health impairment after release of the recovered from hospitals. This has clear economic implications. One hopes that the ICMR’s data set, when released will answer such immediately important questions.

## II. METHODS

The crowd-sourced data set for India [16] used in this study is extracted from state bulletins. The analysis uses 37816 records which contain age information out of a total of 73240 collected till 19 June, 2020. Of these 37110 records contain both age and sex, and 584 of the cases result in fatalities, giving a nominal case fatality ratio of 1.54%.

The national policy for testing during the period covered by this data was largely based on tracing contacts of cases exhibiting severe influenza-like symptoms, with some variation from state to state. As a result, the data set contains cases showing a range of severity, from mild to critical. The incompleteness of the data due to the testing policy could affect the conclusions only if it was biased by age or sex. There are no reports that this is the case.

Since this is a sub-sample of all possible cases which occurred in this period, synthetic samples from the same population of cases can be generated by bootstrap resampling of the base data set. The study of the age and sex distribution of cases involved a 100-fold generation of synthetic data sets of 35000 cases. The study of fatalities is based on 100-fold generation of sets of 500 fatalities. The bootstrap results are given as medians and the 95% confidence limits (CL).

## III. RESULTS

The age distribution of COVID-19 infections marginalized over sex is shown in Figure 1. For comparison, the age distribution from the 2011 census is also shown. Apart from the first two age bins (up to age 19), infections fall faster than the population. Past census data indicate a slight flattening of the age distribution over the years. Correcting for the years passed since the last census is therefore likely to mildly flatten the distribution further. As a result, there is a clear difference between the age distribution of COVID-19 infections and the general population. Younger members of the population are more likely to be infected than a model with constant attack rate would predict. Further work is needed to disentangle the effects of differential attack rate and susceptibility to the infection.

**FIG. 1:**
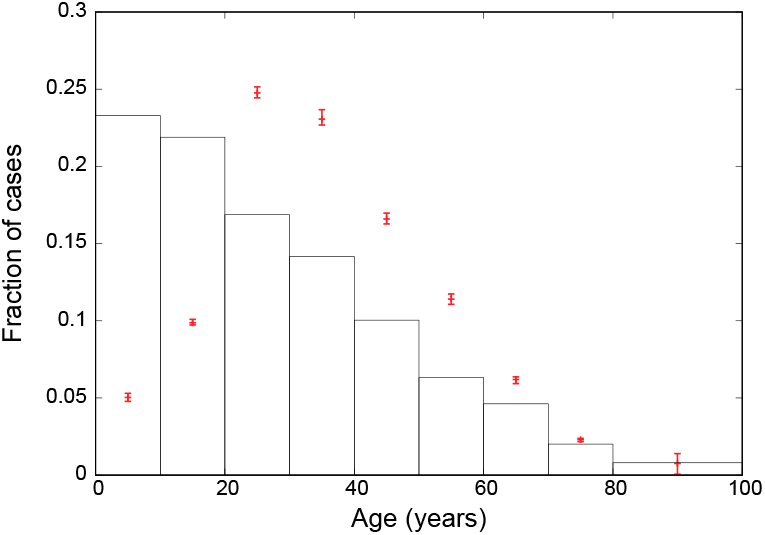
The observed age-distribution of cases marginalized over sexes (boxes, the error bars are the 95% CL from bootstrap resampling). The histogram is the age distribution from the 2011 census.

The age-stratified sex difference in infections is shown in Figure 2. Marginalized over age the female/male ratio of COVID-19 infections is 0.49 with 95% CL in the range [0.48,0.50]. Significant age structuring in the sex difference is absent. Between puberty and menopause the female/male ratio dips below the average. Before puberty, the ratio is closer to unity. The ratio is also larger after menopause. The effect of sex-differentiated attack rate and susceptibility need to be disentangled.

**FIG. 2:**
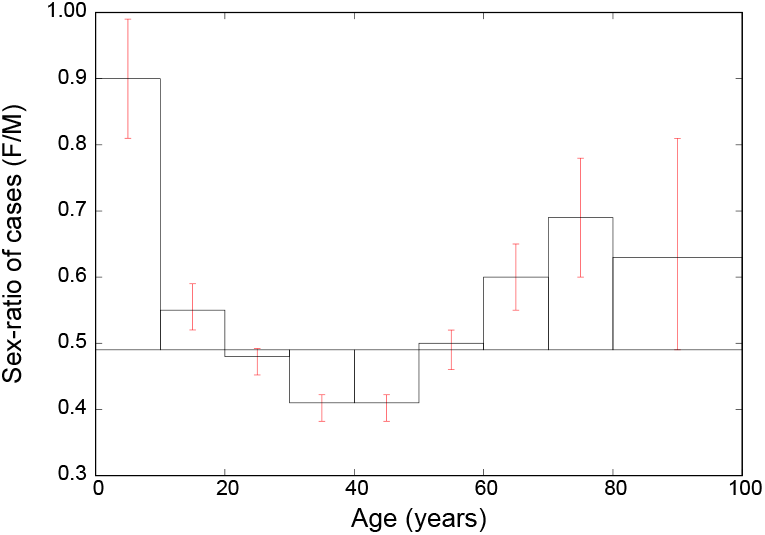
The female/male sex-ratio in observed COVID-19 infections stratified by age. Both the median values (denoted by the histogram) and the error bars denoting 95% CL come from bootstrap resampling of the data. The baseline for the histogram is the median of the distribution marginalized over age.

The above empirical age distribution of infections, is used along with the age stratified IFR in [10] to predict the age distribution of fatalities. The average over the distribution is 0.40% with the 95% CL in the range [0.22%,0.77%]. Since [10] does not provide a sex-differentiated estimate of IFR, these model predictions are blind to sex differences. The comparison of this model with the empirical age distribution of fatalities is shown in Figure 3. There are mild difference between the modal values from the model and the observed distribution, with higher fatalities seen in the 40–59 age group than predicted, and lower fatalities for the age group 70 and above. There is also a puzzling excess of fatalities in the age group 0–9. None of these discrepancies is statistically significant, being within the 95% CrI of the model. It might be argued that the credible interval in each bin is independent of the others, so deviations in multiple bins together may be more significant. Nevertheless, the combined inflluence of the confidence limits on the empirical observations, and the distributions in the model is that there is no significance to differences between the model and data.

**FIG. 3:**
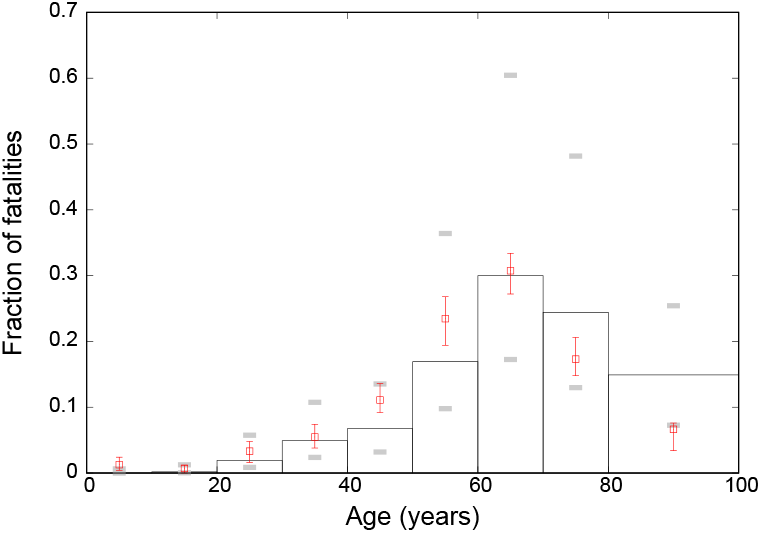
Predictions for the age distribution of fatalities using the observed age distribution of cases along with the age dependence of IFR from [10] (histogram, grey horizontal bars above and below indicate the 95% CrI on each value independently). Also shown is the age distribution of observed fatalities (boxes, the error bars are the 95% CL from bootstrap resampling).

Sex differences are also observed in the fatalities. The age averaged female to male ratio of fatalities is 0.49 with 95% CL [0.41,0.58]. Due to the substantially smaller sample of fatalities, as compared to cases of infection, the age distribution was divided into three categories: ages 0–13, 14–46, and 47 and above. For Indian women, these divisions correspond to (integer parts of) the median ages of puberty and menopause. The female to male ratio for COVID-19 infections and fatalities in these three age bins is given in Table I. All the ratios are significantly smaller than unity, but the age dependence of the ratios for fatalities cannot be seen to have any age structure.

**TABLE I:** The female to male sex ratio of COVID-19 infections and fatalities grouped into three broad age groups, corresponding to the average onset of puberty and menopause in Indian women. The median and 95% confidence limits of the data are shown. For comparison, the sex ratio in the population of India was 0.943 [11].

| Age band | Infections | Fatalities |
| --- | --- | --- |
| 0–13 | 0.53 [0.51,0.56] | 0.5 [0.4, 0.6] |
| 14–46 | 0.44 [0.43,0.45] | 0.5 [0.3, 0.7] |
| 47+ | 0.85 [0.75,0.95] | 0.1 [0,0.6] |

If there is a sex bias in testing which is not dependent on the severity of the case, then it will affect the sex ratio of infections and fatalities equally. A large bias of this kind is therefore unlikely. If the bias depends on the severity of the case, then it is likely that less severe cases are not tested equitably. The postponement of testing until the case develops in severity would lead to a sex difference in the time from disease discovery to recovery. In the small sample of cases where this interval is known [16], the sex-differentiated histograms of recovery times are consistent. So if sex bias in testing exists, it does not explain the full effect seen. Smaller distortions cannot be ruled out in the absence of data on the age and sex distribution of tests administered.

## IV. REMARKS

The age distribution of COVID-19 infections in India is biased towards younger adults more than the hypothesis of uniform attack rate and susceptibility across the whole population would predict. Separating the roles of spatially non-uniform epidemic seeding, age stratified attack rates, co-morbidities, and other factors, needs more detailed data.

This observation can be combined with age stratified IFR [10] to model the age distribution of fatalities. Such a model is in agreement with observation. Since the observation is conditioned on the distribution of infections, no biases are expected.

It is observed that women are half as likely to be infected by COVID-19 as men. There is an interesting age structure to this ratio, with significantly lower infection rates for women between puberty and menopause. The relative importance of sex-linked disease resistance and differential attack rates needs to be clarified with follow up studies. The data for the sex ratio of COVID-19 fatalities does not exhibit any age structure.

## Data Availability

This study only used publicly available data.

https://api.covid19india.org/

## V. DECLARATIONS

Since the data set used is publicly available and anonymized, there are no ethical isuues.

There are no conflicts of interest.

I would like to acknowledge discussion and criticism in the Indian Scientists’ Response to Covid (ISRC) discussion forum. I would also like to thank R. Shankar for a question that led to the design of this study.

